# Deuterium metabolic imaging for assessing response to chemoradiotherapy in high grade glioma: a multisite study

**DOI:** 10.1101/2025.09.30.25336979

**Authors:** Alixander S Khan, Otso Arponen, Giorgia Carnicelli, Ines Horvat-Menih, Nikolaj Bøgh, Maria Jesus Zamora Morales, Esben Søvsø Szocska Hansen, Ashley Grimmer, Elizabeth Latimer, Nichlas Vous Christensen, Michael Vaeggemose, Uffe Kjærgaard, Sebastian Bauer, Jonathan Birchall, Joshua D Kaggie, Rolf F Schulte, Anders Vittrup, Ane B Iversen, Matthew Locke, Marta Wylot, Fiona Harris, Tomasz Matys, Raj Jena, Slávka Lukacova, Mary McLean, Christoffer Laustsen, Ferdia A Gallagher

## Abstract

**Background:** The early assessment of successful treatment response in high-grade glioma is challenging using conventional MRI, due to phenomena such as pseudoprogression which can mimic tumor progression. New methods are needed to evaluate therapeutic efficacy rapidly and accurately. Deuterium Metabolic Imaging (DMI) is a novel, non-invasive technique that can map downstream glucose metabolism in vivo. This study aimed to evaluate the utility of DMI for assessing metabolic response to chemoradiotherapy (ChRT) in glioma patients.

**Methods:** In this prospective two-site study, 18 patients with high-grade glioma underwent 3 T DMI before and after ChRT. Following oral administration of [6,6’-^2^H_2_]glucose, 3D metabolic maps of ^2^H-glucose (Gluc), ^2^H-lactate (Lac), and ^2^H-glutamate+glutamine (Glx) were acquired. Ratios of Lac/Gluc and Glx/Gluc were calculated in the tumor and normal-appearing brain parenchyma to assess glycolytic and oxidative metabolism, respectively.

**Results:** Before treatment, the tumor Glx/Gluc ratio was significantly lower than in the contralateral brain tissue. Following ChRT, a significant decrease in the normalized Lac/Gluc ratio was observed within the tumor volume, indicating a reduction in glycolysis. An important finding was that a lower pre-treatment normalized Glx/Gluc ratio in the tumor was a significant predictor of shorter progression-free survival (median 98 vs. 351 days, p=0.034).

**Conclusions:** DMI identified metabolic changes in high-grade gliomas responding to ChRT. Reduced oxidative metabolism pre-treatment was associated with a poorer prognosis, while post-treatment decreases in glycolysis were demonstrated following therapy. DMI is a promising tool for non-invasively stratifying patients and monitoring treatment efficacy.

## Introduction

High-grade gliomas are the most common primary malignant tumors in the central nervous system (CNS), glioblastoma being the most frequent type and representing up to 45% of all malignant primary brain tumors^1^. The standard of care treatment for grade 4 gliomas in adult patients with a good performance status consists of surgery followed by concurrent chemoradiotherapy (ChRT; radiotherapy (RT) and chemotherapy with temozolomide (TMZ)) followed by 6-12 cycles of maintenance TMZ^2,3^. However, survival times remain poor with the median survival time in glioblastoma (GBM) being only 14.6 months^2^ and the 5-year survival rate is currently only 5.0−9.8%^1,4^. Conventional magnetic resonance imaging (MRI) approaches are typically used to evaluate the morphological response to treatment over time. However, reliable early response evaluation remains challenging due to post-radiotherapy effects resulting in the appearance of an apparent disease progression, including tumor enlargement and increased contrast enhancement, despite the fact the tumor is responding successfully to treatment which is termed pseudoprogression^5^. Alba et al. suggested that nearly half (48.5%) of patients with GBM exhibit tumor enlargement one month after ChRT, with pseudoprogression accounting for up to two thirds, and early true progression for one third of these patients^6^. Distinguishing patients who have early progression and are not responding to ChRT, would allow clinicians to discontinue TMZ and consider experimental therapies. As patients with early true progression cannot be reliably distinguished from patients with pseudoprogression, the former may receive TMZ with no benefit despite potential side effects and may not be considered for experimental therapies until late in their disease course. Indeed, new methods to measure early response are vital to guide treatment decisions at an earlier timepoint and to personalize intervention.

Advanced imaging modalities offer the possibility of non-invasively distinguishing these two groups and facilitating personalized treatment. Multiparametric MRI techniques such as the combined assessment of dynamic contrast enhanced (DCE) and diffusion-weighted imaging (DWI) have improved the assessment of post-ChRT changes^7,8^, but cannot accurately distinguish pseudoprogression since reduced diffusion can be caused by inflammation as well as highly cellular tumor regions^9^. Metabolic imaging has the potential for treatment response evaluation as it exploits the increase in glycolytic metabolism within high grade gliomas. Indeed, taking advantage of a key hallmark of cancer allows for assessment of biological changes before structural changes can be measured. ^18^F-fluorodeoxyglucose positron emission tomography (^18^F-FDG-PET) is widely used as a metabolic imaging tool in clinical practice, but the application of this method in the brain has been limited by the significant normal cerebral uptake of the tracer which makes it difficult to distinguish a tumor from the background brain parenchyma^10^. As an alternative, ^11^C-methionine (MET) PET can be used for molecular targeting of the high uptake of methionine in GBM, and when applied in the post-surgical and post-chemoradiotherapy settings, it has shown the ability to identify response in GBM^11^, but the prognostic information offered by MET-PET is limited^12^. [^18^F]Fluoroethyl-tyrosine (FET) PET offers an alternative PET approach to measure amino acid metabolism within GBM and has shown prognostic value following surgery^13,14^. MR spectroscopy has also been shown high accuracy for detecting true progression by probing steady state metabolite levels^15^, but cannot easily assess metabolic flux information.

Several emerging metabolic MRI techniques have recently provided the ability to non-invasively image downstream intracellular glucose metabolism, and the balance between oxidative and non-oxidative metabolism, rather than glucose uptake alone. Deuterium Metabolic Imaging (DMI) is one such MRI tool where orally administered [6,6’-^2^H_2_] glucose is used to probe metabolic conversion^16^. Spectroscopic approaches can be used with DMI to detect downstream metabolites such as ^2^H-Glx (a combined peak of ^2^H-glutamate and ^2^H-glutamine) and ^2^H-lactate, offering insights into both oxidative phosphorylation in the tricarboxylic acid (TCA) cycle and reductive glycolytic pathways, respectively. The observed ^2^H-glucose offers an indication of the total pool of glucose within the brain and the ratios of ^2^H-Glx or ^2^H-lactate to ^2^H-glucose can be used as an indication of the amount of glucose undergoing oxidative phosphorylation and aerobic glycolysis. DMI has been applied to both the healthy brain^17^ and cerebral diseases such as Alzheimer’s disease^18^ and brain tumors^16,19^. In primary brain tumors, reduced ^2^H-Glx and increased ^2^H-lactate have been observed, reflecting the Warburg effect^16,19^. However, these studies have been affected by the heterogeneity of the disease and are limited to assessment at a single timepoint^16,19^. In this study we have measured metabolic changes of DMI in patients with high-grade gliomas pre- and post-ChRT at two sites, to investigate its potential role in measuring treatment related changes and the correlation with patient outcome.

## Methods

### Patient Recruitment and follow-up

This prospective study performed at two sites (Cambridge University Hospitals NHS Foundation Trust, Cambridge, UK; and Aarhus University Hospital, Aarhus, Denmark) from January 2023 to March 2025, was approved by the institutional research ethics committee in both countries (Ethics Committee of Central Denmark: 1-10-72-118-22, South Central Oxford B Research Ethics Committee: IRAS Number: 294963).

Patients newly diagnosed with high-grade glioma (glioblastoma, GBM, n = 16; high-grade glioma, n = 2; or astrocytoma IDH mutant grade 4, n = 1) scheduled for ChRT were approached and recruited into the study upon written consent. Inclusion criteria required patients to be over 18 years of age and willing to adhere to the use of contraception over the course of the study period if of child-bearing potential. We excluded patients with a contraindication or the inability to tolerate MRI or gadolinium-based contrast agents or high blood glucose level (>10 mmol/L) per the exclusion criteria. Pre-treatment imaging was defined as imaging prior to ChRT following surgical intervention and post-treatment imaging was aimed to occur within 4 weeks of the completion of ChRT. Response was defined by the RANO criteria^5,20^ found during routine clinical follow-up scans.

### Deuterium Metabolic Imaging (DMI)

DMI was performed on 19 patients using two clinical 3 T MRI systems (GE MR 750 (GE Healthcare, Waukesha, WI, USA) for patients 1−14, and GE SIGNA Premier (GE Healthcare, Waukesha, WI, USA) for Cambridge patient 15−19). [6,6’-^2^H_2_]Glucose was administered orally at both sites (doses approved by local ethics committee: Cambridge: 0.75 g/kg body weight; maximum dose, 60 g; Aarhus: 75 g to all patients) after at least 4 h of fasting. Three-dimensional magnetic resonance spectroscopic imaging (MRSI) was acquired at both sites at a mean time of 82 min after glucose administration. Patient 1−6: FOV = 24 cm, matrix size = 10 × 10 × 10, TR = 155.8 ms, flip angle = 70°, number of excitations = 1678, spectral points = 700, bandwidth = 5000 Hz, NEX = 4, total scan time = 17:25 mins, Dual-Tuned ^1^H/^2^H coil (Pulseteq, Cobham, Surrey, UK). Patient 8−14: field of-view FOV = 40 cm, matrix = 16 × 16 × 16, TR = 150 ms, flip angle = 55°, bandwidth = 5000 Hz, number of excitations = 2405, spectral points = 600 points, NEX = 4, total scan time = 24 mins, in-house built DMI coil^17^. Patient 15−19: FOV = 30 cm, matrix = 10 × 10 × 10, TR = 250 ms, flip angle = 60°, number of excitations = 1678, spectral points = 700, bandwidth = 5000 Hz, NEX = 4, total scan time = 28 min, ^2^H-birdcage head coil (Pulseteq, Cobham, Surrey, UK). Patient 7 had the same DMI parameters as patients 8−14 except with FOV = 32 cm and NEX = 2. Prior to DMI measurements, radiofrequency calibration was undertaken to determine the transmit power and resonance frequency and SPGR proton images acquired using the body coil. Following DMI, the patients were temporarily removed from the scanner for head coil exchange, then re-positioned for acquisition of contrast-enhanced T1-weighted images and FLAIR imaging using a 32 channel proton head coil (GE HealthCare). Proton imaging details are provided in the Supplementary Materials. Ten patients from the Cambridge University Hospital cohort also underwent arterial spin labelling (ASL) imaging (labelling pulse duration: 1525 ms (patient 1−11), 2025 ms (patient 12); TR/TE: 4894/10.6 ms; FOV = 24 cm; NEX = 3; slice thickness = 4 mm)

### Image Analysis

Anatomical proton BRAVO images were used to register T1W+Gd images using FSL-FLIRT^21^. Subsequently, regions of interest (ROI) were drawn by two clinical radiology researchers, each with at least 4 years’ experience and reviewed by one another, defining the gross tumor volume as areas of T1W+C enhancement. The contrast enhancing lesion was dilated by 1.8 cm (15 mm clinical target volume + 3 mm isotropic expansion) and the ipsilateral mask applied to ensure the extended region did not cross the midline for pre-treatment and post-treatment evaluation to represent the RT target coverage area. ROIs were created for the following regions: contrast-enhancing lesion + resection cavity (Gross Total Volume, GTV), RT target volume (1.8 cm beyond the GTV region: Planning Target Volume, PTV), Tumor only region (GTV – Resection Cavity), the microscopic disease region (MD: PTV – GTV), ipsilateral normal-appearing brain parenchyma (NABP) and contralateral NABP. Ipsilateral NABP was determined by subtracting the GTV from the ipsilateral hemisphere ROI (Supplementary Materials: Figure 1).

DMI spectra were fitted using OXSA-AMARES^22^ following tMPPCA denoising^23^, partial volume correction^24^ and MICO bias-field correction^25^ using a custom-built MATLAB script. Voxels were rejected prior to fitting if the signal-to-noise ratio (SNR) was <5. Fitting was constrained to have a maximum linewidth of 30 Hz for all the deuterated peaks (water (HDO), ^2^H-Glucose, ^2^H-Glx and ^2^H-Lactate+Lipid) to maintain the accuracy of the measured concentrations. The resulting maps of deuterated water, glucose, lactate, and Glx were interpolated to the resolution of the anatomical images and metabolic values were extracted from the different ROIs. Ratio values were subsequently found from the mean metabolic values for each ROI.

T1W+Gd images were segmented using SPM12^26^ to create brain tissue masks for grey matter (GM) and white matter (WM) which were subsequently divided into contralateral and ipsilateral hemispheres using the drawn ROIs. ASL was registered to the proton images taken with the DMI coil, normalized using the average signal across the whole brain and then values were extracted from the same ROIs as the DMI.

### Statistical Analysis

All statistical analyses were performed using GraphPad Prism (GraphPad Software, San Diego, CA, USA v10.4.1). A p-value <0.05 was considered significant.

Spatial metabolic differences between brain regions of interest (ROIs) before and after ChRT were assessed using ANOVA for multiple comparisons of ROIs to determine significance. To account for global variability, longitudinal analysis was performed using DMI ratios that were first normalized to the contralateral normal-appearing brain parenchyma (NABP). A two-way repeated measures ANOVA was used to assess changes in metabolic metrics across regions and after treatment.

Progression-free survival (PFS) and overall survival (OS) were analyzed using the Kaplan-Meier method with the Gehan-Breslow-Wilcoxon test for group comparisons based on median splits of DMI predictors.

## Results

### Patient Characteristics

A total of 19 patients with glioma grade 4 were recruited for this study across two sites. One patient (Patient 7) was excluded from all DMI analyses due to poor data quality due to variance acquisition parameters, resulting in a final cohort of 18 patients (12 from Cambridge University Hospital, 6 patients from Aarhus University Hospital, age [mean ± standard deviation] of 60 ± 11 years, 17 males and 1 female). Post ChRT scanning was performed 72 ± 11 days after pre-treatment imaging. All patients underwent Chemo-RT (dose 60 Gy in 2 Gy fractions) + adjuvant TMZ except patient 16 who underwent only radiotherapy (40/2.67 Gy) and patients 9 and 12 who passed away before treatment began. Patient characteristics and clinical information are shown in Table 1. One patient (Patient 2) with a midline tumor was excluded from all analyses requiring normalization to the contralateral hemisphere due to the absence of a distinct, uninvolved contralateral region. Tumor volume did not change significantly between the two timepoints (pre-treatment: 22.3 ± 21.2 cm^3^; post-treatment: 18.8 ± 27.8 cm^3^, p = 0.43, Supplementary Materials: Figure 2). ASL was performed in 12 patients pre- and post-treatment, all at Cambridge University Hospital. No significant correlation was found between ASL perfusion and DMI metabolic ratios within the contralateral hemisphere NABP with the closest being Gluc/HDO (p = 0.38, r^2^ = 0.08; Supplementary Materials: Figure 3).

**Table 1.** Summary of clinical information for the 19 patients included in the study.

|  |  |  |  |  |  |  |  |  |  |  |
| --- | --- | --- | --- | --- | --- | --- | --- | --- | --- | --- |
| **Patient number** | **Days between first scan and start of ChRT** | **Days between end of ChRT and second scan** | **EOR** | **Histopathology** | **Location** | **Side** | **IDH status** | **MGMT status** | **Tumor only volume pre-treatment (cm<sup>3</sup>)** | **Tumor only volume post-treatment (cm<sup>3</sup>)** |
| 1 | 5 | 22 | PR | GBM | Frontal | R | 1 | 0 | 4.38 | 4.25 |
| 2 | 6 | 25 | B | GBM | Thalamus | M | 1 | 0 | 50.46 | 77.04 |
| 3 | 1 | 21 | GTR | GBM | Frontal | L | 1 | 0 | 13.51 | 2.50 |
| 4 | 6 | 22 | B | GBM | Temporal | L | 1 | N/A | 2.20 | 0.58 |
| 5 | 3 | 21 | GTR | GBM | Frontal | L | 1 | 1 | 5.16 | 2.61 |
| 6 | 8 | 15 | PR | GBM | Temporal | L | 1 | 1 | 18.43 | 22.04 |
| 8 | 17 | 25 | GTR | GBM | Frontal | R | 1 | 1 | 83.96 | 98.49 |
| 9 | N/A | N/A | GTR | Astrocytoma grade IV | Postero-temporal | R | 0 | 1 | 40.18 | N/A |
| 10 | 17 | 0 | GTR | GBM | Fronto-temporal | R | 1 | 0 | 14.55 | 8.69 |
| 11 | 17 | 25 | GTR | GBM | Parietal | R | 1 | 1 | 9.53 | 4.65 |
| 12 | N/A | N/A | GTR | GBM | Parietal | L | 1 | 1 | 8.36 | N/A |
| 13 | 59 | 22 | B | GBM | Frontal | R | 1 | 1 | 16.61 | 16.56 |
| 14 | 0 | 46 | PR | GBM | Temporal | L | 1 | 1 | 38.01 | 12.40 |
| 15 | 14 | 33 | GTR | HGG | Parietal | R | 1 | 1 | 35.22 | 6.55 |
| 16 | 24 | 26 | GTR | GBM | Frontal | R | 1 | 0 | 10.70 | 10.11 |
| 17 | 62 | 32 | GTR | GBM | Temporal | R | 1 | 0 | N/A | 12.67 |
| 18 | 24 | 26 | GTR | HGG | Medio-temporal | R | 1 | 0 | 16.53 | 14.63 |
| 19 | 21 | 25 | GTR | GBM | Temporal | R | 1 | 0 | 11.09 | 6.50 |
| **Abbreviations.** ChRT = chemoradiotherapy, EOR = Extent of Resection: GTR = gross total resection, PR = partial resection, B = biopsy, Tumor side: R = right, L = left, M = midline, IDH = iso-citrate dehydrogenase: 1 = wild-type, 0 = mutant, MGMT = O<sup>6</sup>-methylguanine-DNA methyltransferase: methylated = 1, unmethylated = 0, GBM = glioblastoma, HGG = high-grade glioma, N/A = not applicable. |  |  |  |  |  |  |  |  |  |  |

#### Metabolic differences between grey and white matter and between hemispheres

At both pre- and post-treatment timepoints, the Lac/Gluc ratio was significantly higher in grey matter (GM) compared to white matter (WM) in both the contralateral (GM: 0.10 ± 0.05; WM: 0.087 ± 0.04) and ipsilateral hemispheres (GM: 0.10 0.047; WM: 0.078 ± 0.045; Figure 1). Metabolic asymmetry was observed for Glx/Gluc between normal appearing grey matter at pre-treatment between the contralateral NABP and ipsilateral NABP. Following treatment, this significant difference in Glx/Gluc measurements between contralateral and ipsilateral grey matter was lost.

**Figure 1.**
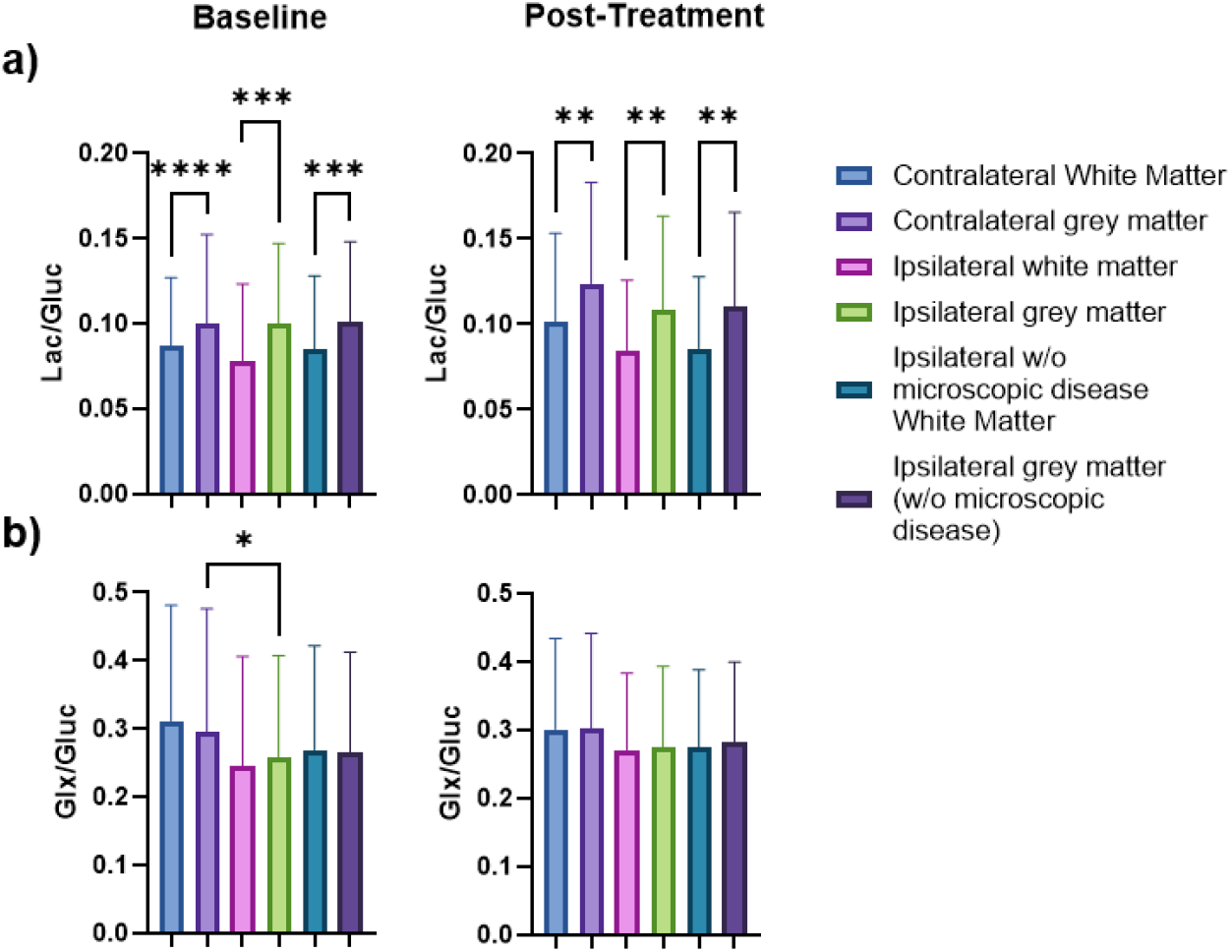
Tissue-specific and hemispheric differences in normal-appearing brain parenchyma. (A) At both baseline pre- and post-treatment time points, grey matter (GM) shows a significantly higher Lac/Gluc than white matter (WM) in both hemispheres, consistent with higher glycolytic activity. (B) At baseline, a significant difference was observed between the hemispheres, with Glx/Gluc being lower in the ipsilateral normal-appearing GM without microscopic disease (NABP GM w/o MD) compared to the contralateral NABP GM, an effect that was not present post-treatment. *, p<0.05, **, p < 0.01, ***, p < 0.001, ****, p < 0.0001, MD = microscopic disease.

#### Spatial variation in metabolism pre- and post-treatment

Spatial differences at both timepoints were assessed between tumor-related regions and the contralateral NABP at both pre- and post-treatment timepoints (Figure 2). At the pre-treatment timepoint, one-way ANOVA showed that Glx/Gluc was significantly higher in the contralateral NABP (0.31 ± 0.17) compared to the tumor only region (0.22 ± 0.15, p = 0.023), the GTV (0.24 ± 0.16, p = 0.02), the ipsilateral hemisphere (0.26 ± 0.15, p = 0.028) and the ipsilateral hemisphere without the microscopic disease (0.26 ± 0.15, p = 0.045). Whilst no significant spatial differences were observed for the Lac/Gluc ratio at pre-treatment timepoint, trends for an increased level within the tumor regions were observed.

**Figure 2.**
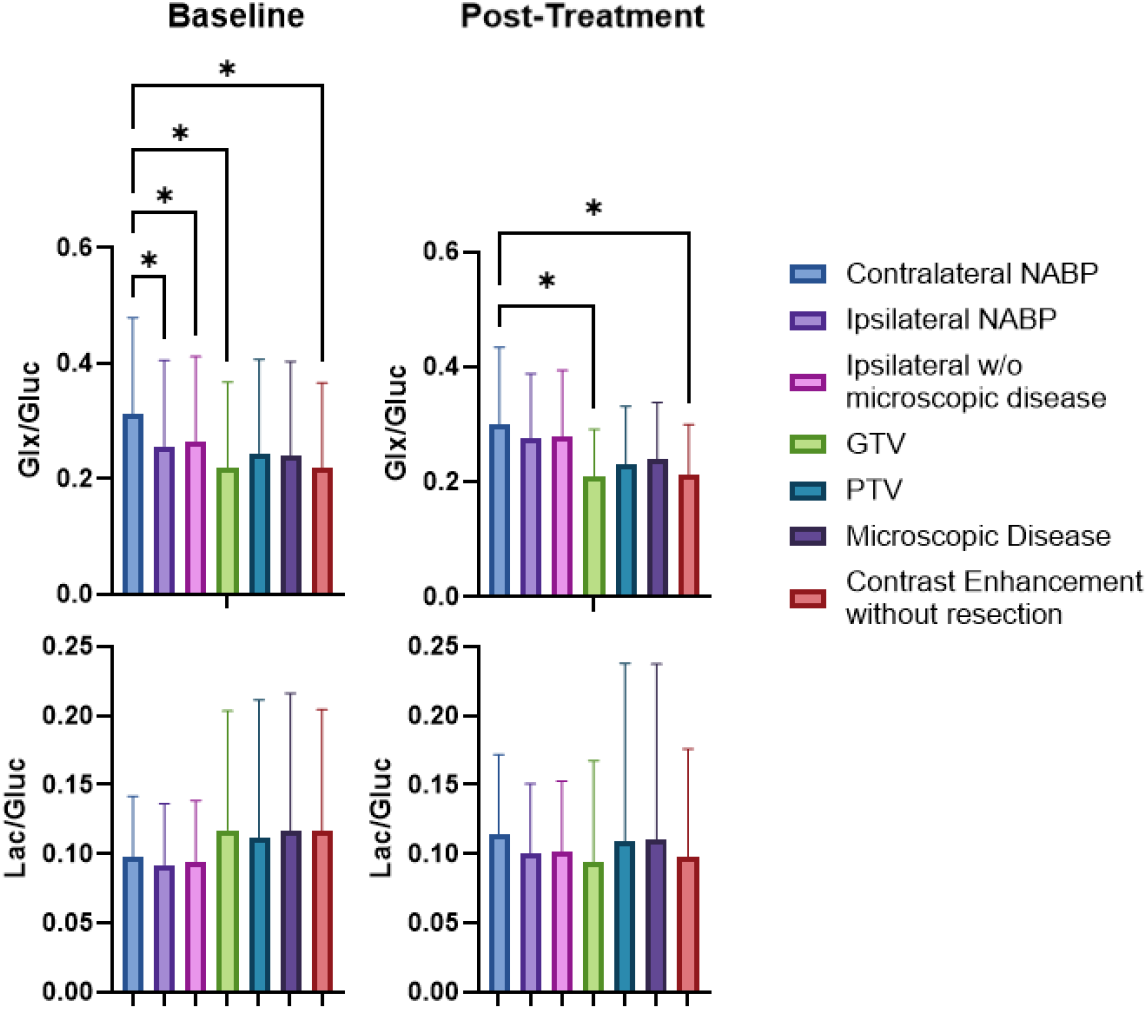
Spatial metabolic differences pre- and post-treatment. Mean ± SD of Glx/Gluc and Lac/Gluc ratios across defined regions of interest. Glx/Gluc was significantly lower in tumor GTV and tumor only regions, as well as the ipsilateral NABP, compared to contralateral NABP at the pretreatment timepoint. These differences between the contralateral NABP and the tumor regions remained significant for Glx/Gluc following treatment, however the significant difference between the ipsilateral hemisphere was no longer present. Whilst no significant differences were seen with Lac/Gluc, increased levels were seen at pre-treatment that removed following treatment.

Following ChRT, Glx/Gluc was higher in the contralateral NABP (0.30 ± 0.13) compared to both the tumor only region (0.21 ± 0.08, p = 0.016) and the GTV (0.21 ± 0.08, p = 0.02), indicating a persistent metabolic abnormality within the residual enhancing tumor. However, significant differences between the contralateral (0.30 ± 0.13) and ipsilateral NABP (0.27 ± 0.12, p = 0.35) were no longer seen. As in the pre-treatment scan, significant differences in Lac/Gluc between ROIs were not seen following treatment.

Figure 3 displays representative pre- and post-ChRT DMI data from patient 18. At the pre-ChRT timepoint, metabolic ratio maps reveal spatial heterogeneity within and around the tumor. The Glx/Gluc map showed lower values within the gross tumor volume (GTV, red arrow) compared to surrounding and contralateral brain tissue (white arrow). Following ChRT, Glx/Gluc maps show a more uniform distribution. Extracted spectra show a lower Glx peak within the GTV compared to the contralateral NABP (fitted spectra shown in Supplementary Materials: Figure 4).

**Figure 3.**
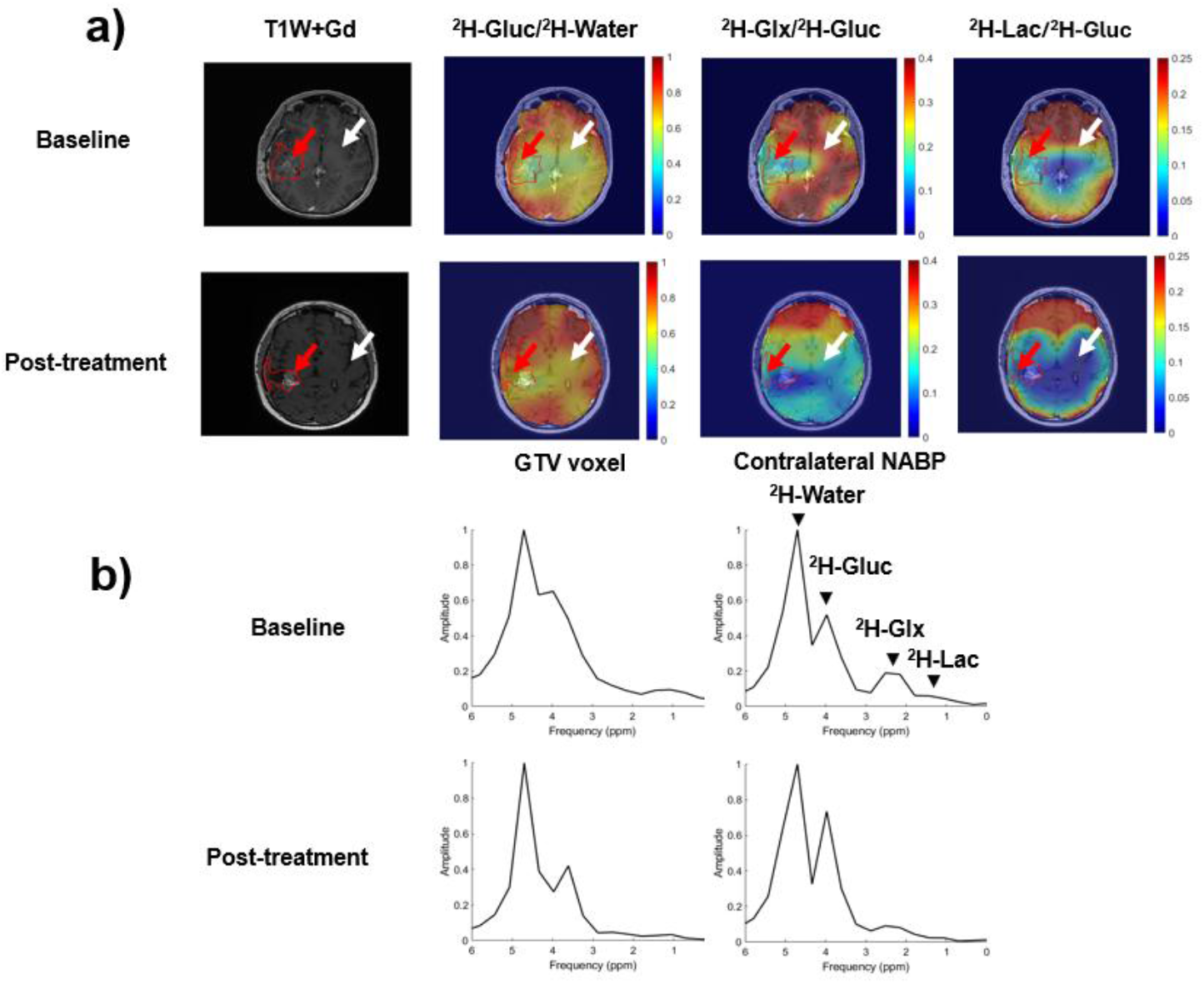
(a) T1W+Gd images and interpolated DMI ratio maps of patient 18 show the tumor (GTV, red ROI) pre-treatment and post-treatment. (b) Corresponding deuterium spectra from the GTV and contralateral NABP show metabolic differences between tumor and healthy-appearing tissue and demonstrate changes within the tumor following therapy.

#### Metabolic changes on DMI following ChRT

To assess longitudinal changes, metabolic values from four ROIs (PTV, MD, tumor only, and ipsilateral NABP) were normalized to the contralateral NABP to account for both technical and physiological differences between patients. These were then assessed across patient cohorts and within individual patients for response (Figure 4). While the glucose uptake (measured as the normalized ratio between glucose and HDO: Gluc/HDO) and aerobic metabolism (measured by Glx/Gluc) remained largely stable before and after treatment across all analyzed ROIs, two-way repeated measures ANOVA on Lac/Gluc showed a significant interaction. Whilst region specific differences of Lac/Gluc were not seen (p = 0.78), treatment effects were significant (p = 0.0018), as well as a significant interaction between the regions and treatment (p = 0.0004). Multiple comparison analysis revealed significant differences in Lac/Gluc ratio following treatment in the PTV (pre-treatment: 1.3 ± 0.85; post-treatment: 0.89 ± 0.80; p = 0.0069) and tumor only region (pre-treatment: 1.4 ± 0.89; post-treatment = 0.82 ± 0.46; p <0.0001). Line plots of the ratio of lactate production to the total glucose pool (Lac/Gluc) and the ratio of Glx production to the total glucose pool (Glx/Gluc) in the PTV and tumor only regions, normalized to the contralateral hemisphere, show individual patient changes. Whilst Lac/Gluc showed an overall group level change within both ROIs, Glx/Gluc showed a heterogeneous response across the cohort.

**Figure 4.**
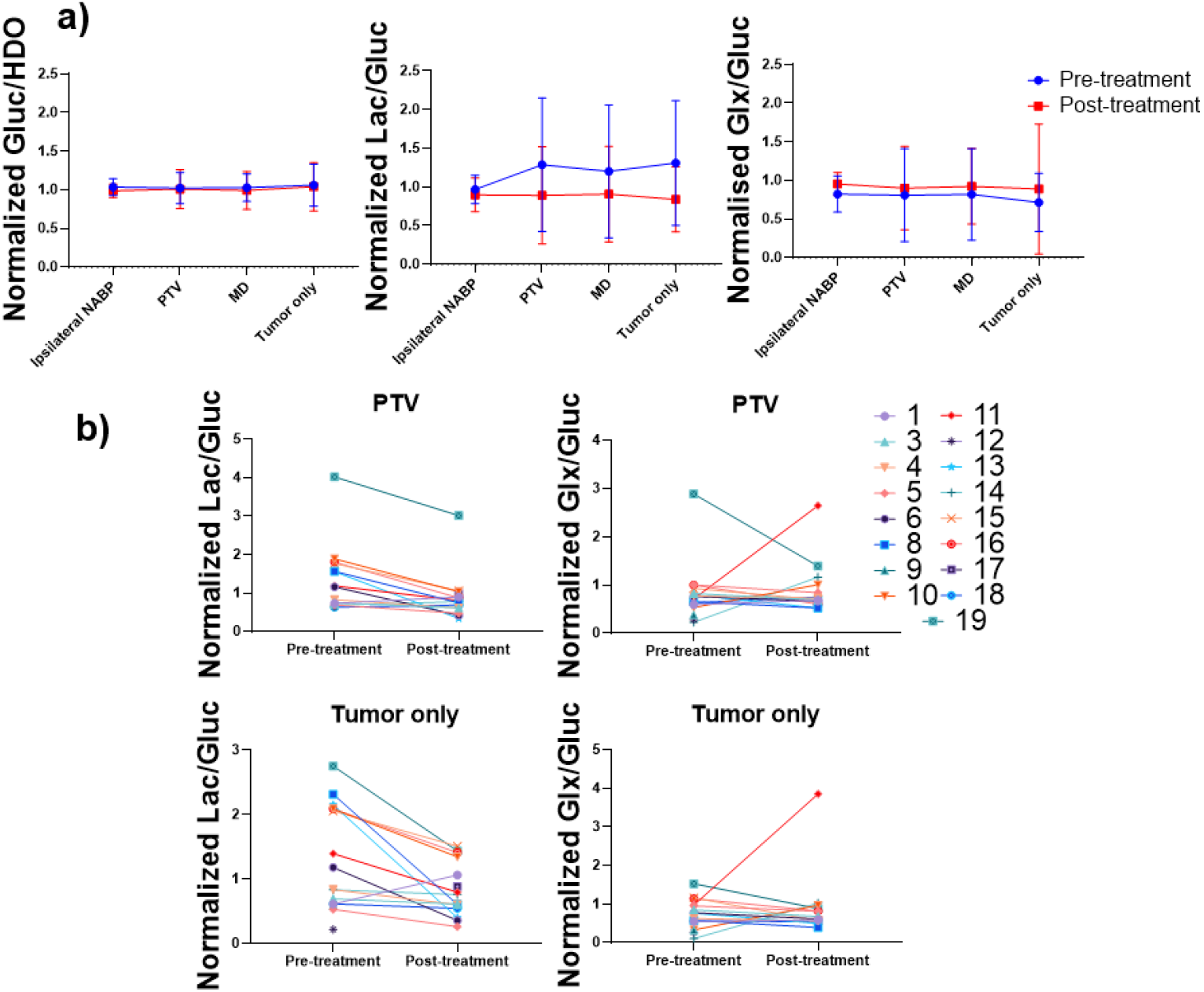
Longitudinal changes in metabolism measured pre- and post-treatment. (a) Means ± standard deviations of normalized ratios at pre-treatment and post-treatment timepoints show a decrease in Lac/Gluc within tumor regions normalized to the contralateral hemisphere but no significant changes of Glx/Gluc or Gluc/HDO. (b) Individual patient trajectories for normalized Lac/Gluc and Glx/Gluc within the planning target volume (PTV) and contrast-enhancing only (Tumor only) regions. The consistent downward trend in the Lac/Gluc line plots illustrates a common metabolic response to treatment, whereas the Glx/Gluc plots highlight a more heterogeneous response profile.

#### Association between metabolism and clinical outcome

A Kaplan-Meier analysis revealed that the pre-treatment Glx/Gluc ratio within the tumor only region, normalized to contralateral NABP, was a significant predictor of patient outcomes (Figure 5). Patients with a pre-treatment Glx/Gluc ratio below the median, had a significantly shorter PFS compared to those with a ratio above the median (median PFS: 98 days vs. 351 days, p = 0.034). While the log-rank test showed a non-significant trend (p = 0.13), the significant result from the Gehan-Breslow-Wilcoxon test suggests that a lower baseline normalized Glx/Gluc is particularly associated with a higher risk of early disease progression. This association extended to OS, where patients with a normalized pre-treatment Glx/Gluc below the median showed a strong trend towards poorer outcomes. However, short follow-up data resulted in the survival rate for values above the median could not be calculated (median OS: 405 days vs. undefined for the above-median group, p = 0.0599). The median OS for the above-median group could not be determined as more than half of these patients were still alive at their last follow-up, further highlighting the survival difference between the groups. In contrast, other DMI metrics, including the pre-treatment Lac/Gluc ratio in the tumor only region and the longitudinal change in any DMI ratio, did not show a significant association with either PFS or OS. Patient characteristics of the cohort above and below the median Glx/Gluc value are included in supplementary materials, Table 1.

**Figure 5.**
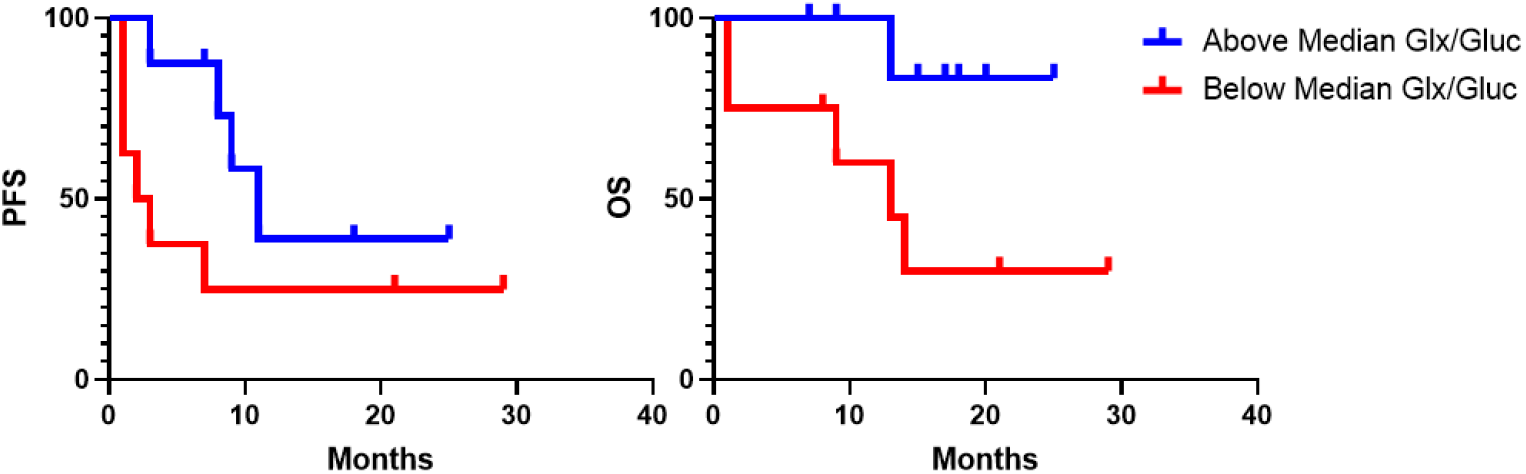
Progression and survival analysis based on metabolic values. Kaplan-Meier analysis shows that patients with a pre-treatment Glx/Gluc ratio below the cohort median within the Tumor only region normalized to the contralateral hemisphere had a significantly worse progression-free survival (PFS, left panel, p = 0.034) and showed a strong trend towards worse overall survival (OS, right panel, p = 0.0599) compared to patients with a ratio above the median.

## Discussion

This study is the first clinical study to demonstrate the use of DMI to measure downstream metabolic changes in high-grade glioma patients in response to ChRT as part of a multi-site study. There was a characteristic metabolic phenotype of a reduction in oxidative metabolism in these tumors, characterized by a significantly lower Glx/Gluc ratio within the contrast enhancing (CE) tumor compared to the contralateral NABP before treatment, as shown previously^16^. This persisted post-ChRT, specifically within the CE region, while peritumoral areas became less distinct from normal tissue. ChRT induced a significant overall decrease in the Lac/Gluc ratio within the broader tumor volume (CE+MD) when normalized to contralateral tissue, indicating a metabolic response to treatment. In comparison, Glx/Gluc did not show any significant longitudinal changes, in part due to the heterogenous directions of the change, lower pretreatment Glx/Gluc ratio pre-ChRT associated with shorter PFS and trended towards shorter OS. Understanding this metabolic heterogeneity in response to treatment in future work may yield important insights into the complex biology of this disease and how it could be harnessed for predicting and detecting response to therapy.

Measurement of metabolic differences between grey and white matter, with elevated levels of ^2^H-lactate seen within the grey matter was used to validate the sensitivity of DMI to measure physiological metabolism. We investigated the correlation to ASL as a measure of perfusion and found no significant correlation across the entire cohort within the contralateral brain between ASL and any DMI metric. This finding indicates that over the timeframe of the DMI experiment using oral administration, glucose delivery is not rate-limited by perfusion, contrasting with alternative metabolic imaging techniques like HP ^13^C-MRI where perfusion can be critical for substrate delivery^17^.

Our observation of altered metabolism in high-grade glioma aligns with a large body of literature demonstrating the Warburg effect and other metabolic reprogramming in tumors^27^. Metabolic reprogramming remains challenging to probe non-invasively with routine clinical imaging. While ^18^F-FDG-PET has shown increased glucose uptake in high-grade glioma, its utility is limited by high background uptake in the brain^10^. Amino acid PET with ^18^F-FET and ^11^C-MET offer an alternative^11^, but DMI provides a unique window into downstream glucose metabolism. Emerging metabolic imaging tools offer potential methods to assess tumor metabolism and offer potential for early diagnosis^28,29^ and insights into identifying early and successful treatment response^30,31^. The potential clinical utility of DMI has been demonstrated in the brain^16,19,29,32,33^ and in other areas of the body^34^. Previous applications of DMI in brain tumors^16,19^ have been in small patient populations and at single timepoints with limited analysis of metabolic variation across the brain. Our results show that DMI can detect significant metabolic differences between the areas of residual tumor and the NABP both pre-treatment and following therapy. Significant reductions in ^2^H-Glx/Gluc within both the tumor region and the ipsilateral NABP compared to the contralateral NABP indicated the expected metabolic shift away from oxidative citric acid cycle metabolism towards aerobic glycolysis. Grey matter differences between hemispheres pre-treatment also demonstrated a significant reduction in ^2^H-Glx/Gluc within the ipsilateral hemisphere indicating impairment of oxidative metabolism within the grey matter but not the white matter. This difference may be because of hydrostatic pressure effects or tumor inflammation arising in the ipsilateral hemisphere. The metabolic alterations between the ipsilateral and contralateral hemispheres are concordant with the highly infiltrative nature of high-grade glioma, where despite surgical debulking there is microscopic disease adjacent to the resected tumor. This finding demonstrates the challenge in managing patients with high grade glioma where the tumor often extends beyond the contrast enhancing region^35,36^.

Longitudinal measurements of metabolic changes found a decrease in glycolysis as represented by the ^2^H-Lac/Gluc ratio within the PTV region normalized to the contralateral hemisphere following ChRT. The measured decreased demonstrates that DMI has the sensitivity to measure treatment related changes that could be utilized for measurement of early treatment response where conventional imaging is often confounded by pseudoprogression in up to a third of patients after ChRT^5,6,9^. The results here suggest that DMI could provide a more direct biological measure of treatment effects, potentially helping to distinguish true progression from treatment-related changes earlier than anatomical imaging alone, and offers a potential avenue for future research. Decreases in the ^2^H-Lac/Gluc may occur due to several treatment-related effects, including a selective reduction in glycolytic tumor cells, a shift in the metabolism of remaining cells away from aerobic glycolysis and towards oxidative phosphorylation^37^, or improved oxygenation within the tumor microenvironment following the radiotherapy^38^. This reduction in the relative lactate pool suggests that ChRT successfully modulates the Warburg phenotype that characterizes high grade glioma.

Finally, our survival analysis revealed that the pre-treatment normalized ^2^H-Glx/Gluc ratio below the median within the tumor only region was a significant predictor of poorer PFS and showed a strong trend towards worse OS. This novel and potentially clinically important finding suggests that the degree of metabolic reprogramming away from oxidative metabolism in the tumor core before treatment, as indicated by a lower Glx/Gluc ratio, could be a powerful prognostic marker. Tumors exhibiting a more pronounced Warburg-like phenotype pre-treatment may represent a more aggressive disease^39^. Pre-clinical studies that have applied DMI to GBM^40^ have shown the ability of DMI to differentiate metabolic subtypes that relate to prognosis^41^. This measurement may also reflect several factors related to surgical outcome: for instance, tumors with a greater burden of metabolically active residual cells following incomplete resection would likely present an altered metabolic profile and are also known to be associated with poorer outcomes^42,43^. Our finding provides a metabolic correlation to this clinical observation and suggests pre-treatment DMI could offer a non-invasive method for assessing the metabolic burden of residual disease post-surgery.

The results suggest that DMI could address several unmet challenges in neuro-oncology imaging. Differentiating true tumor progression from treatment-related effects such as pseudoprogression, radionecrosis^44^, and pseudoresponse^45^, are clinically challenging as all these can mimic tumor recurrence on conventional MRI^46^. Given that DMI provides a direct readout of metabolic activity independent of vascular changes as shown here, it has the potential to distinguish viable, metabolically active tumor from treatment effects, thereby reducing diagnostic delays and ensuring patients are allocated to appropriate therapies more quickly. Furthermore, DMI could improve the characterization of the peritumoral region and the detection of MR-invisible disease where most recurrences occur^47^. Our observation of a significantly altered Glx/Gluc in the ipsilateral NABP as well as the ipsilateral grey matter regions outside the main tumor mass, provides metabolic evidence of this infiltrative field effect that cannot be detected easily on conventional imaging. DMI can non-invasively map these metabolic alterations and assess tumor heterogeneity beyond the limits of a single-site biopsy, and is a promising tool to complement standard MRI, potentially refining radiotherapy planning and allowing for more personalized patient care in the future.

Our study has several limitations. First, the cohort size is small and the population heterogeneous, which limits the statistical power for survival analyses and subgroup comparisons. However, this is mitigated by the multi-site nature of this study, which is the first such DMI study to date, providing support for the use of the technique to produce meaningful results across a variety of patient populations and centers. Second, there is an absence of corresponding tumor tissue for direct biological correlation of our metabolic findings with genomic or proteomic data due to the nature of the study design. Third, post-surgical inflammation and the effects of steroids can alter overall metabolism and may confound the interpretation of true tumor metabolism^48^. A significant challenge for DMI, particularly at 3T, is the low signal-to-noise ratio and spectral resolution for lactate, as this peak can be contaminated by signal from nearby lipids. As lipid signals are stable over short time periods, any change in this combined peak will represent dynamic changes in lactate labelling which can therefore be deconvolved from the overall peak, but it remains as a technical consideration. In addition to spectral challenges, the resolution of DMI is limited, which can lead to partial volume effects, potentially blurring the metabolic distinctions between ROIs. Whilst preprocessing of the DMI data aims to reduce blurring with partial volume correction and denoising, further work to improve overall spatial resolution to measure intratumoral heterogeneity is needed. Finally, whilst this two-site study demonstrates repeatability and provides external validity to the findings, the different parameters used in scanning could impact results. Most notably, different TR and flip angles used as part of the MRSI may affect sensitivity to lactate and water due to their longer T_1_ (however, all patients underwent the same scanning parameters at baseline and post-treatment to mitigate this). Relaxation times of deuterated metabolites have not yet been reported in human brain at 3T, and further work is needed to find the optimal MR weighting for accurate and reproducible indications. Despite these limitations, the repeatability of DMI at clinical field strengths has been previously reported to have a coefficient of variation (CoV) of 10% within subjects^49^ and the successful application of the technique across two clinical sites here further supports its potential for wider use in multi-center studies.

In summary, this multi-site longitudinal DMI study demonstrates that metabolic signatures can differentiate high-grade glioma from NABP and that these signatures evolve in response to ChRT. Specifically, we identified pre-treatment normalized Glx/Gluc in the tumor only region as a significant prognostic biomarker for PFS, linking a more pronounced Warburg-like phenotype to poorer outcomes. Furthermore, we demonstrated the longitudinal decrease in normalized Lac/Gluc within the CE+MD region as a promising biomarker of treatment response. Despite current technical limitations, these findings highlight the potential of DMI to provide non-invasive, biologically relevant information for patient stratification and treatment response assessment in high-grade glioma.

## Acknowledgments

We thank Tau Vendelboe for imaging assistance.

## Data availability

The data that support the findings of this study are available from the corresponding author upon reasonable request.

