## Supplementary Materials for "Deuterium metabolic imaging for assessing response to chemoradiotherapy in high grade glioma: a multisite study"

**Methods: Proton Imaging sequences**

SPGR with body for DMI anatomical localisation:

Patients 1-6: inversion time = 450 ms; FOV = 240 mm; TR = 5.4 ms; echo time (TE) = 1.7 ms; flip angle (FA) = 12°; spatial resolution = 0.9 × 0.9 × 1 $mm^3$

Patient 7-12: inversion time = 450 ms; FOV = 240 mm; TR = 7.4 ms; echo time (TE) = 3.1 ms; flip angle (FA) = 12°; spatial resolution = 0.9 × 0.9 × 2 $mm^3$

Patient 13-19: inversion time = 600 ms; FOV = 240 mm; TR = 7.4 ms; echo time (TE) = 3.1 ms; flip angle (FA) = 12°; spatial resolution = 0.5 × 0.5 × 2 $mm^3$

T1W +C:

Patients 1-6: FOV =256; TR = 6.2 ms; echo time (TE) = 2.7 ms; flip angle (FA) = 12°; spatial resolution = 0.9 × 0.9 ×0.5 $mm^3$

Patient 7-12: FOV = 256 mm; TR = 8.2 ms; echo time (TE) = 3.2 ms; flip angle (FA) = 90°; spatial resolution = 1 × 1× 1 $mm^3$

Patient 13-19: FOV = 256 mm; TR = 4.8 ms; echo time (TE) = 11.9 ms; flip angle (FA) =90°; spatial resolution = 0.5 × 0.5 × 0.8 $mm^3$

FLAIR:

Patients 1-6: FOV = 256 mm; TR = 5.4 ms; echo time (TE) = 123 ms; flip angle (FA) = 160°; spatial resolution = 0.9 × 0.9 × 4 $mm^3$

Patient 7-12: FOV = 256 mm; TR = 6202 ms; echo time (TE) = 103 ms; flip angle (FA) = 90°; spatial resolution = 0.4 × 0.4 × 1 $mm^3$

Patient 13-19: FOV = 256 mm; TR = 6000 ms; echo time (TE) = 122 ms; flip angle (FA) =90°; spatial resolution = 0.5 × 0.5 × 0.8 $mm^3$

**Figure 1.** Tumour ROIs.

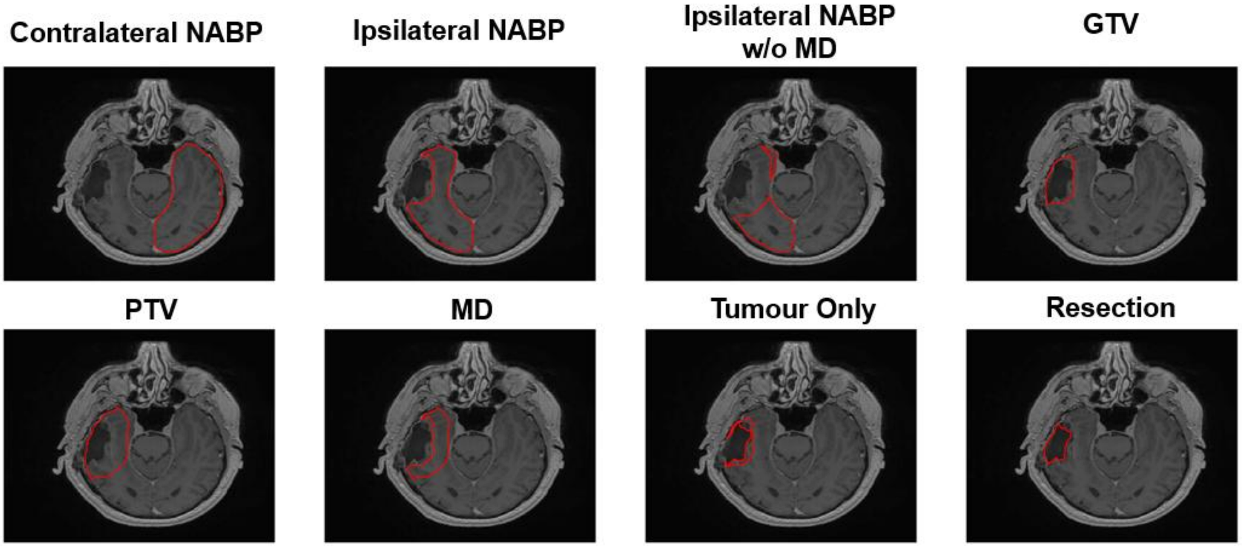


ROI = region of interest, NABP = normal-appearing brain parenchyma, GTV = gross tumour volume, PTV = planning target volume MD = microscopic disease.

**Figure 2**. Tumour volume changes.

Pre-treatment = 22.29 ± 21.19 $cm^3$
Post-Treatment = 18.77 ± 27.81 $cm^3$
Unpaired t-test p = 0.42

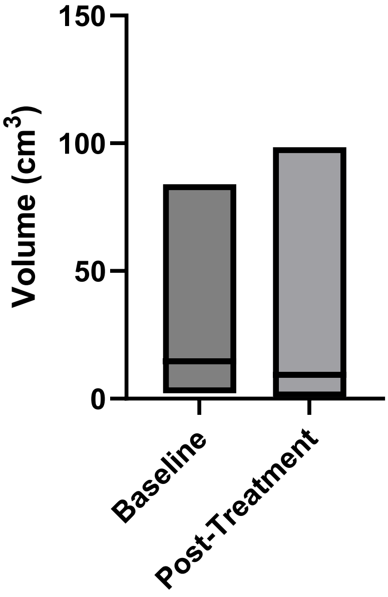


**Figure 3.** ASL and DMI correlation analysis

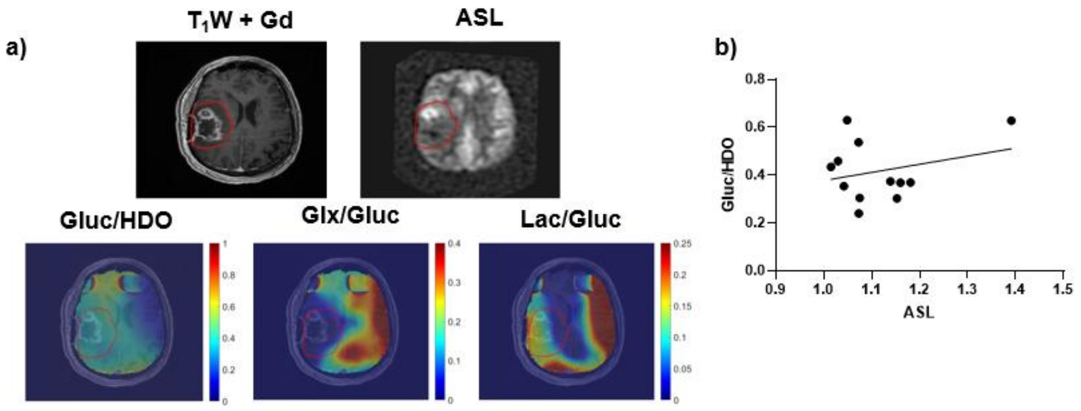


**ASL and DMI correlation:** (a) Representative anatomical, perfusion (ASL), and metabolic (DMI) maps. (b) A scatter plot of ASL against vs. Glu/HDO values ratios from the contralateral normal-appearing brain parenchyma (n = 12 scans). No significant correlation was found between ASL perfusion and any DMI metabolic ratios, indicating they provide independent physiological information.

**Figure 4**. OXSA AMARES peak fitting on tumour only and contralateral NABP region

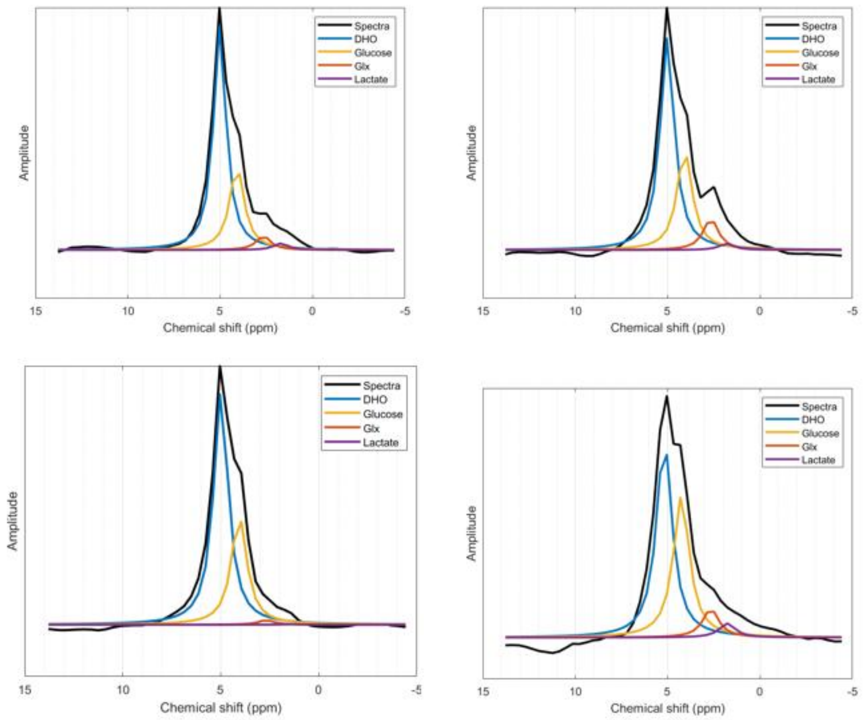


**Table 1.** Patient characteristics of above and below median Glx/Gluc within the tumour only region.

**Table 1 – Table of patient characteristics above and below the median Glx/Gluc measured at baseline**

**Below Median**

**Extent of Resection: GTR = gross total resection, PR = partial resection, B = biopsy, Tumor side: R = right, L = left, M = midline, IDH = iso-citrate dehydrogenase: 1 = wild-type, 0 = mutant, MGMT = O$^6$-methylguanine-DNA methyltransferase: methylated = 1, unmethylated = 0**

| Patient number | Days between first scan and ChRT | Days between end of ChRT and second scan | Extent of resection | Tumour side | IDH | MGMT | Tumour only volume pre-treatment (cm$^3$) | Tumour only volume post-treatment (cm$^3$) |
|---|---|---|---|---|---|---|---|---|
| 1 | 5 | 22 | PR | R | 1 | 0 | 4.38 | 4.25 |
| 4 | 6 | 22 | B | L | 1 | N/A | 2.20 | 0.58 |
| 8 | 17 | 25 | GTR | R | 1 | 1 | 83.96 | 98.49 |
| 9 | N/A | N/A | GTR | R | 0 | 1 | 40.18 | N/A |
| 10 | 17 | 0 | GTR | R | 1 | 0 | 14.55 | 8.69 |
| 12 | N/A | N/A | GTR | L | 1 | 1 | 8.36 | N/A |
| 14 | 0 | 46 | PR | L | 1 | 1 | 38.01 | 12.40 |
| 18 | 24 | 26 | GTR | R | 1 | 0 | 16.53 | 14.63 |

**Above Medianss**

| Patient number | Days between first scan and ChRT | Days between end of ChRT and second scan | Extent of Resection | Tumour Side | IDH | MGMT | TO volume pre-treatment ($cm^3$) | TO volume Post-treatment ($cm^3$) |
|---|---|---|---|---|---|---|---|---|
| 3 | 1 | 21 | GTR | L | 1 | 0 | 13.51 | 2.50 |
| 5 | 3 | 21 | GTR | L | 1 | 1 | 5.16 | 2.61 |
| 6 | 8 | 15 | PR | L | 1 | 1 | 18.43 | 22.04 |
| 11 | 17 | 25 | GTR | R | 1 | 1 | 9.53 | 4.65 |
| 13 | 59 | 22 | B | R | 1 | 1 | 16.61 | 16.56 |
| 15 | 14 | 33 | GTR | R | 1 | 1 | 35.22 | 6.55 |
| 16 | 24 | 26 | GTR | R | 1 | 0 | 10.70 | 10.11 |
| 19 | 21 | 25 | GTR | R | 1 | 0 | 11.09 | 6.50 |
